# Stroke Incidence According to Cardiorespiratory Fitness: A Cohort Study of 483,379 Hypertensive Patients

**DOI:** 10.1101/2023.10.06.23296681

**Authors:** Peter Kokkinos, Charles Faselis, Andreas Pittaras, Immanuel Babu Henry Samuel, Carl J. Lavie, Robert Ross, Michael Lamonte, Barry A. Franklin, Xuemei Sui, Jonathan Myers

## Abstract

**Objectives:** We assessed stroke incidence in hypertensive patients according to cardiorespiratory fitness (CRF) and changes in CRF.

**Methods:** A prospective cohort study of 483,379 US Veterans. Participants completed a maximal standardized Exercise Treadmill test (ETT) performed within the Veterans Affairs medical centers across the United States between 1999 and 2020. None exhibited evidence of unstable cardiovascular disease during the ETT. Participants were stratified into 5 age-and-gender specific CRF categories based on the peak metabolic equivalents (METs) achieved. A subgroup of participants with two ETT evaluations (n=110, 576) were also assigned to 4 categories based on MET changes from the initial ETT to the final ETT. Multivariable Cox models, adjusted for age, and co-morbidities were used to estimate HRs and 95% CIs for stroke risk.

**Results:** The mean age ± standard deviation (SD) was 59.4±9.0 years. During the median follow-up time of 10.6 years (5,182,179 person-years), there were 15,925 stroke events with an average annual rate of 3.1 events per 1,000 person-years. In a final adjusted model, relatively poor CRF was the strongest predictor of stroke risk than any other comorbidity (HR: 2.24; 95% CI: 2.10-2.40; P< 0.001). For each 1-MET higher exercise capacity, the risk was 10% lower (0.90, 95% CI 0.90-0.91, p<0.001). Compared to the Least-fit, stroke risk was 23% lower for Low-fit individuals (HR 0.77; 95% CI, 0.73-0.80; p<0.001); and declined progressively to 55% for those in the highest CRF category (HR 0.45; 95% CI 0.42-0.48; p<0.001). We also assessed stroke incidence according to change in CRF. Compared to fit individuals during both evaluations, the risk was 27% higher for those who became unfit (HR 1.27, 95% CI 1.15-1.41, p<0.001), and not significantly different for unfit who became fit (HR 1.10, 95% CI 0.97-1.25, p=0.13).

**Conclusions:** Poor CRF was the strongest predictor of stroke incidence in hypertensive patients, regardless of age race, or gender. The association was independent, inverse, and graded for all stroke types. Changes in CRF over time reflected inverse changes in stroke risk, suggesting that risk of stroke can be modulated by improved CRF.

## Introduction

Stroke is the leading cause of long-term disability and the fifth leading cause of death in the United States.^1^ Age-adjusted stroke incidence and stroke-related death rates are higher among African American, Hispanic, and Native American adults compared to non-Hispanic White (White) adults,^1,2^ despite recent efforts to reduce racial disparities in stroke prevention and treatment.^1,3^ High blood pressure and diabetes are considered the major risk factors for stroke and contribute to increased risk among African American adults with higher prevalence of both health conditions. ^1, 2, 4–6^

Physical inactivity is one of ten potentially modifiable risk factors, that account for over 90% of the population-attributable risk for stroke worldwide.^7^ Epidemiologic evidence from large studies strongly supports an inverse and independent association between cardiorespiratory fitness (CRF), assessed objectively using a standardized exercise treadmill test (ETT) expressed in metabolic equivalents (METs; 1 MET=3.5 ml of O_2_ per kg of body weight per minute), and chronic disease incidence, and all-cause mortality. ^8–13^ Self-reported physical activity (PA) status suggests that increased PA is associated with lower stroke incidence and recurrence, ^14–18^ likely the outcome of the favorable effects PA exerts on blood pressure control, lipid and carbohydrate metabolism, and endothelial function.^1^

Studies that have defined CRF objectively by a ETT have also demonstrated an inverse association between CRF and risk for stroke. ^19–23^ Recently, two relatively small studies that examined changes in CRF by sequential standardized ETTs have reported that improvements in CRF are independently and inversely associated with a lower risk of ischemic stroke. ^23, 24^

To our knowledge, the association between the risk of stroke in hypertensive patients and CRF assessed by a standardized ETT, has not been assessed. Furthermore, most of the available evidence in this area is based on studies conducted among predominantly white populations.

There is no information on the CRF-stroke risk association among AA, Hispanic and Native American adults, the most vulnerable populations, and minimal information exists on septuagenarians and octogenarians. Finally, little is known regarding the association between changes in CRF that lead to reciprocal changes in stroke risk for hypertensive patients.

Thus, the main objective of the current study was to assess stroke incidence in hypertensive US Veterans according to CRF defined by a standardized ETT across race and age categories. An additional objective was to evaluate the association between stroke risk and changes in CRF, assessed by a two standardized ETTs obtained at least one year apart. We hypothesized that the association between CRF status and stroke incidence would be inverse and graded, regardless of race, or age. We also hypothesized that changes in CRF would reflect changes in the incident rate of stroke. The Veterans Affairs Health Care System maintains a nationwide extensive electronic health care database. This, and the equal access to medical care independent of a patient’s financial status, provides a unique opportunity to determine health outcomes more precisely and completely compared with other cohorts, ^25^ while minimizing the influence of medical care disparities. ^26–28^

## METHODS

### Study Population

The cohort of the current study consisted of 483,379 US Veterans (23,171 women). Of these, 348,389 were White (72.1%), 98,761 (20.4%) African American, 23,580 (4.9%) Hispanic, 10,272 (2.1%) American Indian/Pacific Islander/ Alaska Native/Asian; and 2,377 (0.5%) were unknown or declined to disclose. All participants met the following criteria: 1) they were diagnosed with hypertension determined by at least 2 consecutive diagnostic recordings at least 6 months apart; 2) had no diagnosis of stroke prior to hypertension or 6 months post-hypertension; and 3) were followed for at least 6 months post-hypertension (Figure 1).

**Figure 1.**
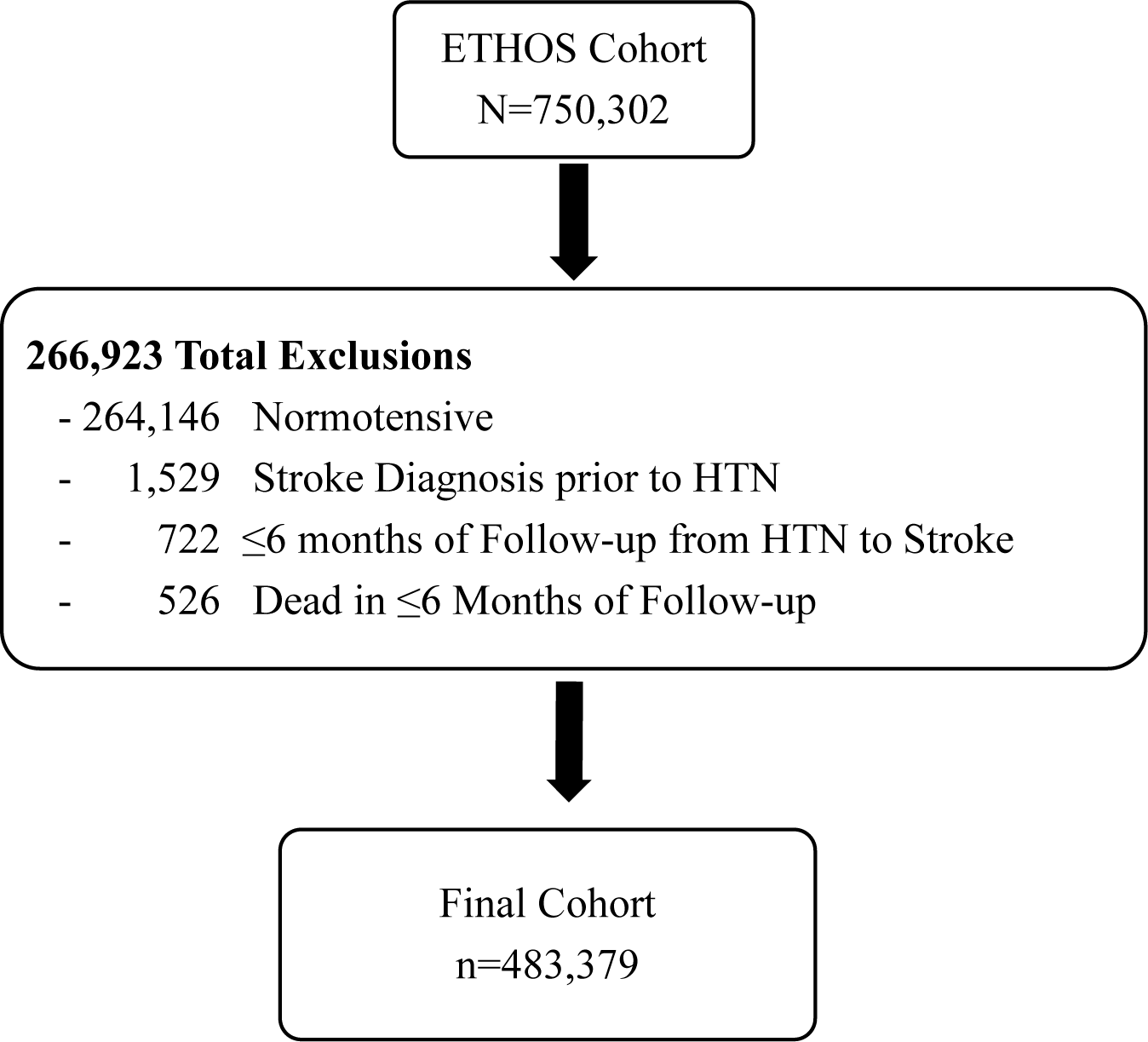
Flow chart for cohort selection. Flowchart depicts exclusions of subjects based on predetermined criteria and the final cohort. HTN= hypertension.

The current cohort was derived from the ongoing Exercise Testing and Health Outcomes Study (ETHOS), a larger cohort (n=750,302), based at the Veterans Affairs Medical Center in Washington, DC. The ETHOS study was initiated in 1999 and included two Veterans Affairs (VA) medical centers located in Washington, DC., and Palo Alto, CA. As electronic records were instituted in all VA medical centers, additional data were collected from VA medical centers nationwide. The selection of the original cohort is described in detail elsewhere. ^9^ Briefly, all participants completed a maximal ETT performed using the Bruce protocol within the Veterans Affairs hospital system across the United States between October 1, 1999, and September 3, 2020. Participants were selected if they were 30 to 95 years of age at the time of ETT, achieved an exercise capacity of 2.0 or more metabolic equivalents of task (METs), exhibited no evidence of unstable CVD during the ETT, had a body mass index (BMI) of 18.5 kg/m^2^ or higher, and had a follow-up period of at least 6 months; those receiving statin therapy were treated for at least 1 year. To lower the likelihood of including individuals with underlying serious heart disease that was not detected during the ETT, we also excluded individuals who met the following conditions within 6 months after ETT: those who underwent coronary artery bypass grafting, had percutaneous transluminal coronary angioplasty or percutaneous coronary intervention, and had a myocardial infarction or a diagnosis of heart failure.

### Procedures

Detailed information on relevant demographic characteristics, clinical and medication information, risk factors, and comorbidities as defined by the International Classification of Diseases, Ninth and Tenth Revision coding, with at least 2 recordings at least 6 months apart, were obtained for all participants from the Veterans Affairs Computerized Patient Record System. The Veterans Affairs records have high sensitivity for identifying the incidence of chronic conditions.^29^ Historical information included the onset of previous myocardial infarction, cardiac procedures, heart failure, hypertension, hypercholesterolemia, diabetes mellitus, renal disease, stroke, cancer (all), smoking status, aspirin, use of statins, hypoglycemics, and cardiac or antihypertensive medications.

Data and analyses are presented in accordance with the Strengthening the Reporting of Observational Studies in Epidemiology reporting guideline for cohort studies.^30^

The study was approved by the institutional review board at the Washington, DC, Veterans Affairs Medical Center (IRB Net# 1602164).

### Extraction of METs

The extraction of peak METs achieved is described in detail elsewhere.^9^ Briefly, we randomly selected 3,000 samples of physician clinical notes on exercise capacity from the data set and identified METs manually. This annotated data set was used to train Natural Language Processing models to recognize the words METs and MET. Once METs were extracted, the MET data were randomly and manually checked for errors. The model accuracy on the test data set was 97%.

### Cardiorespiratory Fitness Categories

Peak METs were calculated for each participant by standardized equations based on treadmill speed and grade.^31^ Age- and gender-specific CRF categories based on METs achieved were established based on methods described in our previous work.^32^ Briefly, we first stratified the cohort into 5 age groups (30 to 49 years, 50 to 59 years, 60 to 69 years, 70 to 79 years, and 80 to 95 years). We then identified the age- and sex-specific MET level that corresponded to 20th, 40th, 60th, and 80th percentiles within their respective age category. Finally, we combined the respective percentiles to form the following 5 CRF categories: least fit (n=92,596; METs= 4.3±1.2), low fit (n=121,154; METs, 6.9±1.1), moderately fit (n=88,844; METs, 8.3±1.2), fit (n=127,271; METs, 10.3±0.9), and highly fit (n=53,514; METs, 13.2±1.6).

### CRF Categories Based on Changes in Exercise Capacity

To evaluate the association between changes in CRF and risk of stroke, we identified 110, 576 patients with two ETT assessments at least one year apart (mean ± SD; 6.0±3.9 years). We then calculated the age-specific median MET level achieved at the initial ETT assessment 8.0 METs and 7.0 METs for ages <60 years and ≥60 years, respectively), and defined those below and above the median as Unfit and Fit, respectively. Individuals defined as Unfit who achieved a MET below this median during the final ETT assessment were reclassified as Unfit/Unfit (n=45,922), and those defined as Unfit at baseline and achieved a MET level above the median were reclassified as Unfit/Fit (n=9,289). Similarly, individuals defined as Fit initially who achieved a MET level above the median during the final ETT assessment were reclassified as Fit/Fit (n=41,638), and those defined as Fit initially and achieved a MET level below the median were reclassified as Fit/Unfit (n=13,727).

### Progression Rate to Stroke

The primary outcome was diagnosis of stroke obtained for all participants from the Veterans Affairs Computerized Patient Record System and verified by at least 2 consecutive recordings at least 6 months apart. International Classification of Diseases, ninth (ICD-9 430 to 438) and tenth Revision (ICD-10 I60 to I69) coding were used to identify diagnosis of stroke.

### Statistical Analyses

Continuous variables are presented as means ± SD and categorical variables as relative frequencies (percent). We tested baseline mean differences of normally distributed variables between individuals within CRF categories with one-way ANOVA and differences between categorical variables with χ^2^ tests. The assumption of equality of variances between groups was tested by Levene’s test and the assumption of normality with probability-probability plots.

We estimated hazard ratios (HRs) and 95% confidence intervals (CI) associated with event rates (stroke incidence) across the CRF categories using Cox proportional hazards models for the entire cohort, and according to age categories, sex, and race. For these models, we used the Least-fit CRF category (20^th^ percentile) as the referent to access the risk associated with increased CRF status and the High-fit category (80^th^ percentile) to access the risk associated with decreased CRF status. We also estimated HRs and 95% CI associated with event rates (stroke incidence) across categories depicting CRF changes. For these models we used both the Unfit/Unfit category as the reference groups to assess the risk associated with increased CRF status and the Fit/Fit category to assess the risk associated with decreased CRF status.

The follow-up time for those who suffered a stroke was calculated from the date of hypertension diagnosis to the first recorded date of stroke. For those who did not develop stroke follow-up time was calculated to the date of death, the last date of VA care, or the end of the follow-up period (September 30, 2021), whichever came first.

All analyses assessing the risk of stroke were adjusted for the following comorbidities assessed prior to the diagnosis of hypertension: age, BMI, race, gender, history of atrial fibrillation, all other CVD, risk factors (diabetes mellitus [T2DM], dyslipidemia, smoking, alcohol abuse, sleep apnea), chronic kidney disease (CKD), and medications. In addition, for the models formed to assess risk according to CRF change we adjusted for the time in years between the two assessments.

The assumption of proportionality for all Cox proportional hazard analyses was tested graphically by plotting the logarithm of cumulative hazards concerning each covariate separately. The proportionality assumption was fulfilled for each model. All hypotheses were two-sided and p<0.05 was deemed statistically significant. All statistical procedures were performed with SPSS (version 29.0).

## Results

### Patient Demographics

The mean age ± standard deviation (SD) of the entire cohort was 59.4±9.0 years. The follow-up time ranged from 0.51 to 22.0 years (mean, 10.7±5.1 years) and a median of 10.6 years (IQR 6.6-14.6 years), providing 5,182,179 person-years of follow-up. There were 15,925 stroke events (3.3%) with an average annual rate of 3.1 events per 1,000 person-years.

Patients who developed stroke were significantly older by approximately 2.0 years (mean age 61.4 ± 8.9 years vs 59.3 ± 8.9 years) and had higher systolic blood pressure (141.6 ± 15.6 mm Hg vs 139.0 ± 14.5 mm Hg). They also had significantly higher prevalence of T2DM (27.3 % vs 20.3%), cardiovascular disease (3.1% vs 2.6%), atrial fibrillation (3.4% vs 2.3%), and chronic kidney disease (3.5% vs 2.6%). The prevalence of smoking (15.8% vs 16.9%), and sleep apnea (6.6% vs 9.5%) was significantly lower in the stroke group.

Demographic and clinical characteristics across the CRF categories are presented in Table 1. Systolic blood pressure (BP), and BMI tended to be lower with increased CRF, with age being slightly lower by approximately one year only in the highest CRF category. Similarly, CVD risk factors and overall disease burden were progressively more favorable for those in the higher CRF categories compared to those in the lowest CRF category.

**Table 1.** Clinical characteristics of the cohort according to CRF categories.

| Variables | Least-fit<br>(n=92,596) | Low-fit<br>(n=121,154) | Moderately-fit<br>(n=88,844) | Fit (n=127,271) | Highly-fit<br>(n=53,514) | p-value |
| --- | --- | --- | --- | --- | --- | --- |
| Race/Ethnicity (%) |  |  |  |  |  |  |
| *Caucasian* | 66,805 (19.2) | 88,794 (25.5) | 63,520 (18.2) | 91,785 (26.3) | 37,893 (10.9) |  |
| *African American* | 19,819 (20.0) | 24,611 (24.9) | 17,899 (18.1) | 25,504 (25.8) | 11,022 (11.1) |  |
| *Hispanic* | 3,967 (16.8) | 4,858 (26.6) | 4,751 (20.1) | 6,694 (28.4) | 3,327 (14.1) |  |
| *Native-American, Alaskan,*<br>*Hawaiians, or Asian* | 1,673 (16.3) | 2,414 (23.5) | 2,317 (22.5) | 2,798 (27.2) | 1,075 (10.5) |  |
| *Unknown* | 488 (20.5) | 615 (25.9) | 435 (18.3) | 601 (25.3) | 240 (10.1) |  |
| Age (yrs) | 59.3±8.8 | 59.6±8.8 | 59.6±9.3 | 59.6±8.6 | 58.4±9.7 | <0.001 |
| Systolic BP (mm Hg) | 140.4±15.3 | 140.0±14.8 | 139.2±14.4 | 138.2±14.1 | 136.6±13.5 | <0.001 |
| Diastolic BP (mm Hg) | 80.7±9.3 | 80.9±9.0 | 80.8±8.8 | 80.9±8.8 | 80.8±8.6 | <0.001 |
| Peak METs achieved | 4.3±1.1 | 6.6±1.1 | 8.1±1.3 | 9.8±1.2 | 12.7±1.2 | <0.001 |
| Body Mass Index (kg/m<sup>2</sup>) | 31.0±6.2 | 30.5±5.4 | 30.0±4.9 | 29.4±4.5 | 28.4±3.9 | <0.001 |
| Smoking Status (%) | 37,750 (40.8) | 42,951 (35.5) | 27,575 (31.0) | 37,139 (29.2) | 13,625 (25.5) | <0.001 |
| Alcohol Abuse (%) | 13,916 (15.0) | 15,420 (12.7) | 10,337 (11.6) | 14,109 (11.1) | 5,801 (10.8) | <0.001 |
| Diabetes Mellitus (%)<sup>a</sup> | 24,452 (26.4) | 28,333 (23.4) | 17,995 (20.2) | 22,196 (17.4) | 6,155 (11.5) | <0.001 |
| Cardiovascular disease (%)<sup>b</sup> | 38,751 (41.8) | 46,461 (38.3) | 32,380 (36.4) | 44,750 (35.2) | 16,353 (30.6) | <0.001 |
| Atrial Fibrillation (%) | 3,827 (4.1) | 4,513 (3.7) | 3,079 (3.5) | 4,069 (3.2) | 1,616 (3.0) | <0.001 |
| Dyslipidemia (%)<sup>c</sup> | 76,176 (82.3) | 98,603 (81.4) | 71,361 (80.3) | 101,750 (79.9) | 40,910 (76.4) | <0.001 |
| Chronic kidney disease (%)<sup>d</sup> | 19,524 (21.1) | 19,769 (16.3) | 12,884 (14.5) | 15,001 (11.8) | 4,811 (9.0) | <0.001 |
| Sleep Apnea (%) | 39,637 (42.8) | 46,924 (38.7) | 34,058 (38.3) | 47,503 (37.3) | 18,900 (35.3) | <0.001 |
| Statins (%) | 73,787 (79.7) | 93,140 (76.9) | 65,592 (73.8) | 91,543 (71.9) | 35,289 (65.9) | <0.001 |
| Hypoglycemics (%) | 43,508 (47.0) | 50,835 (42.0) | 32,511 (36.6) | 40,459 (31.8) | 11,442 (21.4) | <0.001 |
| Cardiac/antihypertensives (%) | 87,491 (94.5) | 111,981 (92.4) | 80,695 (90.8) | 113,288 (89.0) | 45,075 (84.2) | <0.001 |

Data shown as Means ± SD unless specified otherwise.
<sup>a</sup> Diabetes mellitus was defined as consistently elevated glucose levels $\geq 126$ mg/dL, or HbA1c $\geq 6.5\%$ or history of physician-diagnosed diabetes mellitus.
<sup>b</sup> Cardiovascular disease was defined as history of physician-diagnosed MI, coronary artery bypass graft surgery, congestive heart failure, stroke, and peripheral vascular disease.
<sup>c</sup> Dyslipidemia was defined as Total cholesterol $\geq 200$ mg/dL, or LDL-C $\geq 130$ mg/dL or HDL-C $< 40$ mg/dL or a combination.
<sup>d</sup> Chronic kidney disease (CKD) was defined as a history of eGFR $< 60$ mL/min per $1.73 \text{ m}^2$ or physician-diagnosed CKD.

### Predictors of Stroke Risk

In a final adjusted model, the association between CRF and the risk of stroke was inverse and graded. For every 1-MET higher exercise capacity, the final adjusted HR for stroke was 10% lower (0.90, 95% CI 0.90-0.91, p<0.001) for the entire cohort. The rate was similar for men and women, Whites, Blacks and Hispanics, and those aged <70 years. For those ≥70.0 years of age, the risk was 6% lower per 1-MET increase (HR 0.94, 95% CI 0.92-0.95, p<0.001) and for Asian/Native Americans there was a 14% per 1-MET increase in exercise capacity (HR 0.86, 95% 0.82-0.90; p<0.001).

The adjusted risk of stroke associated with CRF categories and clinical comorbidities are presented in Figure 2. Poor CRF emerged as the strongest predictor of risk. Compared to the High-fit category (referent), stroke risk was highest for those in the Least-fit category (HR: 2.24; 95% CI: 2.10-2.40; P< 0.001), followed by Low-fit (HR: 1.72; 95% CI: 1.61-1.84; P< 0.001), Moderate-fit (HR:1.49; CI: 1.39-1.59; p<0.001) and Fit (1.21 (1.13-1.29; p<0.001). Other predictors of incident stroke included: age at the time of HTN onset (per 10 years) HR 1.36, 95% CI 1.33-1.38; p<0.001), CKD (HR 1.71, 95% CI 1.65-1.78; p<0.001), history of atrial fibrillation (HR 1.54, 95% CI 1.44-1.66; p<0.001), dyslipidemia (HR 1.40, 95% CI 1.34-1.46; p<0.001), T2DM (HR 1.33, 95% CI 1.28-1.37; p<0.001), sleep apnea (HR 1.33, 95% CI 1.29-1.38; p<0.001), alcohol abuse (HR 1.21, 95% CI 1.15-1.27; p<0.001) and CVD (HR 1.15, 95% CI 1.11-1.19; p<0.001), and systolic BP (per 10 units), HR 1.09, 95% CI 1.07-1.10; p<0.001).

**Figure 2.**
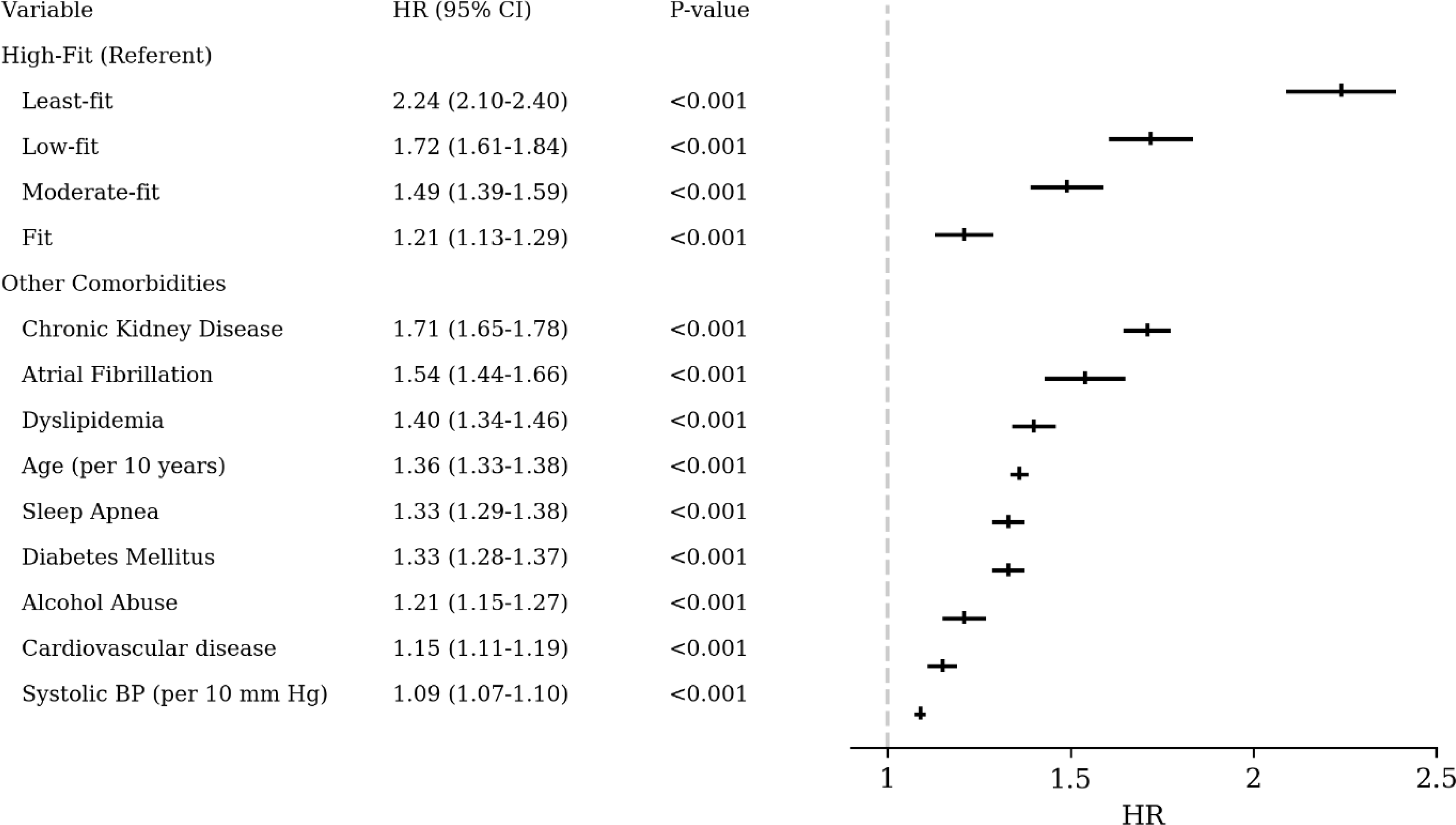
Forest plot of HRs and 95% CIs from multivariable Cox regression analysis examining the association between stroke and select clinical characteristics.

We also examined the risk associated with CRF across the five CRF categories for the entire cohort and subgroups of gender, race, and age using the Least-fit category as the referent. The findings of these analyses are presented in Table 2. We observed clear and strong inverse dose-response gradients for incident stroke across the incremental categories of CRF (overall p<0.001). The stroke risk was 23% lower for the Low-fit (HR 0.77; 95% CI, 0.73-0.80; p<0.001); 34% for Moderately fit (HR 0.66; 95% CI 0.63-0.69; p<0.001); 46% for Fit (HR 0.54; 95% CI, 0.51-0.56; p<0.001), and 55% lower for individuals in the highest CRF category (HR 0.45; 95% CI 0.42-0.48; p<0.001). Similarly, significant inverse trends in stroke risk related to increased CRF were also observed in men, women, for each race/ethnicity group and the three age categories. However, the impact of CRF was somewhat attenuated in those aged 70 years and older.

**Table 2.** Risk of Stroke Across Fitness Categories for the Entire Cohort and Subcategories.

|  | **Least-fit**<br>**(n=92,596)** | **Low-fit**<br>**(n=121,154)** | **Moderately-fit**<br>**(n=88,844)** | **Fit**<br>**(n=127,271)** | **High-fit**<br>**(53,514)** |
| --- | --- | --- | --- | --- | --- |
| Entire Cohort (n=483,379) | 1.0 | 0.77 | 0.66 | 0.54 | 0.45 |
|  | --- | (0.73-0.80) | (0.63-0.69) | (0.51-0.56) | (0.42-0.48) |
| No of Event | 4,267 (4.6) | 4,394 (3.6) | 2,844 (3.2) | 3,305 (2.6) | 1,115 (1.9) |
| *Peak METs (Mean ± SD)* | *4.3±1.1 METs* | *6.6±1.1METs* | *8.1±1.3 METs* | *9.8±1.2 METs* | 12.7±1.7 METs |
| Men (n=460,208) | 1.0 | 0.77 | 0.66 | 0.54 | 0.45 |
|  |  | (0.73-0.80) | (0.63-0.70) | (0.51-0.56) | (0.42-0.49) |
| Women (n=23,171) | 1.0 | 0.76 | 0.64 | 0.56 | 0.47 |
|  |  | (0.61-0.95) | (0.49-0.84) | (0.43-0.74) | (0.28-0.81) |
| Caucasians (n=348,389) | 1.0 | 0.77 | 0.66 | 0.55 | 0.46 |
|  | -- | (0.73-0.81) | (0.62-0.70) | (0.52-0.58) | (0.43-0.50) |
| African-Americans | 1.0 | 0.78 | 0.72 | 0.54 | 0.42 |
| (n=98,761) | -- | (0.71-0.9851) | (0.65-0.79) | (0.49-0.60) | (0.36-0.48) |
| Hispanics (23,580) | 1.0 | 0.70 | 0.64 | 0.49 | 0.43 |
|  | -- | (0.58-0.85) | (0.52-0.78) | (0.40-0.60) | (0.33-0.56) |
| Native-American; Asian; | 1.0 | 0.90 | 0.58 | 0.40 | 0.31 |
| Alaskan/Hawaiians | -- | (0.66-1.22) | (0.41-0.81) | (0.28-0.58) | (0.18-0.56) |
| (n=10,272) |  |  |  |  |  |
| <60 yrs of age; (n=261,331) | 1.0 | 0.73 | 0.60 | 0.50 | 0.40 |
|  | -- | (0.69-0.78) | (0.56-0.65) | (0.47-0.54) | (0.36-0.44) |
| *No of Events/No at Risk (%)* | 2,238/51,454 (4.3) | 2,059/63,795 (3.2) | 1,289/49,313 (2.6) | 1,462/65,952 (2.2) | 502/30,817 (1.6) |
| *Peak METs (Mean ± SD)* | 4.7±1.2 | 7.2±0.9 | 9.0±1.0 | 10.3±0.9 | 13.3±1.4 |
| 60-69 yrs of age; (n=167,040) | 1.0 | 0.78 | 0.66 | 0.53 | 0.45 |
|  | -- | (0.73-0.84) | (0.61-0.72) | (0.49-0.58) | (0.40-0.51) |
| *No of Events/No at Risk (%)* | 1,445/30,927 (4.7) | 1,640/44,032 (3.8) | 894/26,850 (3.3) | 1,239/48,359 (2.6) | 375/16,872 (2.2) |
| *Peak METs (Mean ± SD)* | 4.0±0.8 | 6.3±0.8 | 7.4±0.6 | 9.5±0.9 | 12.2±1.5 |
| 70-95 yrs of age (n=55,008) | 1.0 | 0.87 | 0.85 | 0.72 | 0.61 |
|  | -- | (0.78-0.97) | (0.76-0.95) | (0.64-0.80) | (0.52-0.71) |
| *No of Events/No at Risk (%)* | 584/10,215 (5.7) | 695/13,327 (5.2) | 661/12,681 (5.2) | 604/12,960 (4.7) | 238/5,825 (4.1) |
| *Peak METs (Mean ± SD)* | 3.4±0.7 | 4.9±0.6 | 6.5±0.7 | 7.9±1.1 | 10.5±1.3 |
| Stroke 3-years Post HTN | 1.0 | 0.78 | 0.66 | 0.55 | 0.45 |
| Excluded (n=2,522) | -- | (0.74-0.81) | (0.63-0.70) | (0.52-0.58) | (0.42-0.48) |
| No Stroke-Dead within 3 yrs | 1.0 | 0.76 | 0.66 | 0.53 | 0.44 |
| post-HTN (n=13,525) | -- | (0.73-0.79) | (0.62-0.69) | (0.51-0.56) | (0.41-0.47) |

Finally, we examined the CRF-stroke risk association according to the type of stroke, (ischemic, hemorrhagic, and arachnoid). The inverse and graded association between CRF and stroke risk was evident regardless of the stroke type. The risk was approximately 40%-50% lower for individuals with an exercise capacity of approximately 10.0 METs (Fit category) compared to the Least-fit individuals (Figure 3).

**Figure 3.**
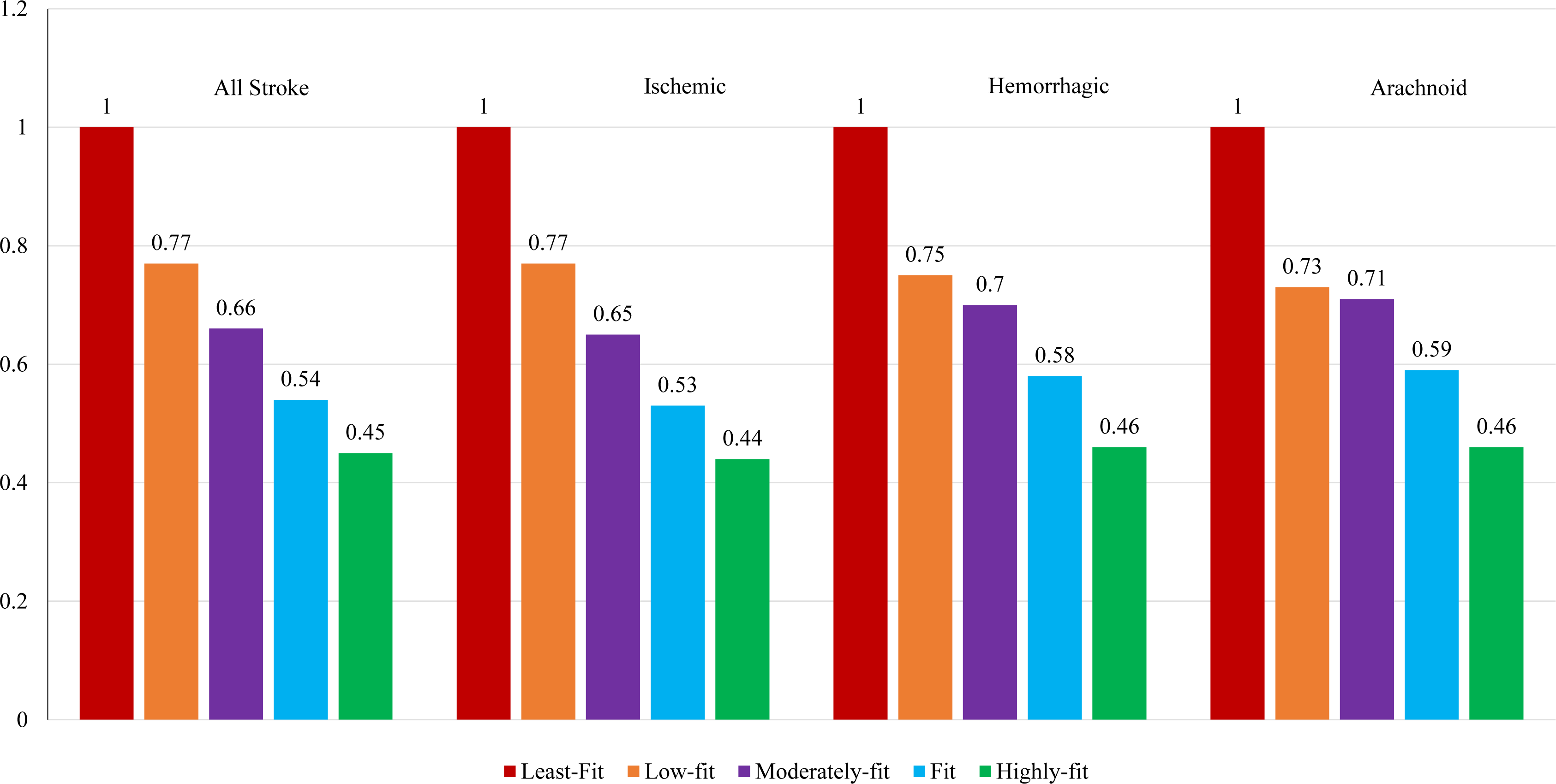
Relative risk of stroke according to cardiorespiratory fitness categories

### Change in CRF and Stroke Incidence

In a subset of our cohort with two CRF assessments (n=110,576) over a mean period of 6.0±3.9 years, we evaluated the impact of change in CRF on the risk of stroke. The proportion of repeat ETT was approximately 22% of individuals within the Least-fit and Low-fit CRF categories, 23% in the Moderate-fit and Fit categories and 24% in the High-fit category.

Individuals who were classified as fit during both assessments (Fit-Fit) served as the reference group. The risk of stroke was 27% higher (HR 1.27, 95% CI 1.15-1.41, p<0.001) for fit individuals who became unfit at the final assessment (Fit-Unfit). For those who were classified as unfit during both evaluations (Unfit-Unfit), the risk of stroke was 55% higher (HR 1.55, 95% CI 1.44-1.67, p<0.001). No significant increase in risk was observed in unfit individuals who became fit (HR 1.10, 95% CI 0.97-1.25, p=0.13; Figure 4).

**Figure 4.**
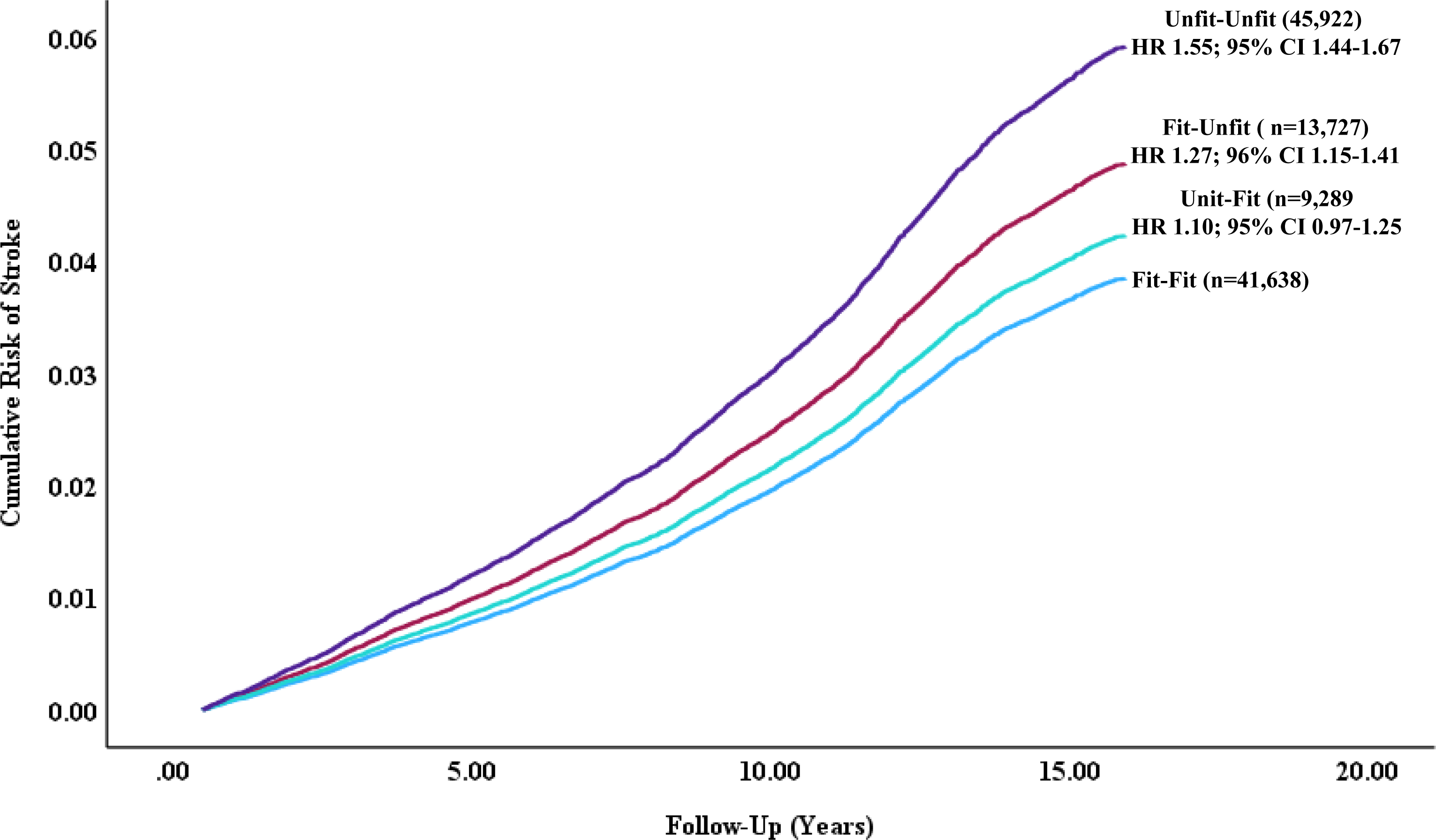
Stroke risk according to changes in cardiorespiratory fitness

We also used the Unfit-Unfit group as the referent to gain perspective on the impact an increase in CRF may have on the risk of stroke in unfit individuals. The risk of stroke among those who were classified as unfit initially but improved their fitness status at the final ETT (Unfit-Fit) was 29% lower (HR 0.71, 95% CI 0.63-0.80, p<0.001).

### Reverse Causality and Competing Risk

To account for the possibility that the higher stroke rates and the decline in CRF were the outcomes of underlying and unrecognized disease (reverse causality) and not poor CRF, we excluded individuals who developed stroke within three years post hypertension (n=2,522). To account for competing risk of progression to stroke, we excluded those who died prior to developing stroke (n=13,525). In both sub-analyses performed after these exclusions, the association between CRF and stroke risk did not change substantially from that observed in the entire cohort (Table 2).

## Discussion

The findings of the current study support an inverse and graded association between CRF and the risk of stroke in hypertensive patients. The association was independent of all traditional risk factors and was similar for men and women, and across the racial and age spectra. It was also true for ischemic, hemorrhagic, and arachnoid strokes.

It is also of clinical and public health importance that significantly lower risk rates were observed with peak CRF of approximately 5.0, 6.0 and 7.0 METs for ages ≥70, 60-69 and <60 years, respectively. These MET levels are achievable by most middle-aged and older individuals by meeting current guidelines that advocate ≥150 minutes per week of moderate or 75 minutes of vigorous physical activity.

Our findings extend the observations of previous, relatively small studies.^19–22^ and make unique contributions to the existing literature. First, this is largest cohort of hypertensive patients (n=483,373) with objectively assessed CRF with extensive data on comorbidities and cardiometabolic medications. The size of our cohort also allowed us to examine the CRF-stroke risk association in the largest cohort of African American (n=98,855), Hispanic (n=23,597), and American Indian/Pacific Islander/ Alaska Native/Asian subjects (n=10,277). Stroke incidence and stroke-related death rates are higher among these groups; ^1, 2, 4^ this despite efforts to reduce racial disparities in stroke prevention and treatment.^1, 3^ Additionally, it is the largest cohort of participants aged ≥70 years (n=55,008) as data in the elderly have been previously either limited or unavailable. Finally, it allowed adequate statistical power to assess the CRF-stroke risk for the specific stroke types.

Previous, relatively small studies and in populations that were not exclusively hypertensive have reported that changes CRF over time were associated with parallel changes in the risk of stroke. ^23, 24^ In the current study, the relatively large sample of patients with two ETT assessments at least one year apart (n=110, 576), allowed a more detailed examination of the association between CRF changes the risk of stroke. We found that changes in CRF reflected inverse and proportional changes in the risk of stroke. Compared to fit individuals during both evaluations (Fit-Fit), the risk was 55% higher for individuals who were unfit during both evaluations, and there was no significant risk for unfit subjects at baseline who became fit.

Conversely, the risk was 27% higher for fit individuals who became unfit. These findings suggest that the risk of stroke can be modulated by improved CRF regardless of initial CRF status. It should also be noted that the CRF-related protection against stroke is diminished if CRF is not maintained. Thus, the importance of improving and/or maintaining fitness should be emphasized by health care professionals.

### Mechanisms

The precise physiological mechanisms involved in the favorable impact of CRF on stroke incidence are beyond the scope of this study. Cumulative evidence from previous studies suggests that the CRF-related protection against stroke incidence may result from the modulatory effects of CRF on several systems. For example, exercise-mediated reduction in resting and exercise blood pressure for hypertensive patients is well established. ^31, 34^ Improved CRF has also been shown to improve endothelial function in hypertensive patients ^35, 36^ and has a favorable impact on several cardiometabolic co-morbidities ^37–40^ associated with increased risk of stroke.^1, 2^ In the current study we observed that the overall disease burden (i.e., blood pressure, diabetes mellitus, CVD, CKD, etc.,) was progressively lower with higher CRF (Table 1). Evidence also supports an association between poor CRF and greater abnormal left ventricular remodeling, diastolic dysfunction, and decreased contractility.^41^ Conversely, improved cardiac function by significantly decreasing cardiac stiffness has been reported in a recent exercise intervention study following 2 years of aerobic exercise training. ^42, 43^ Collectively, these findings support that increased CRF is likely to lessen the disease burden and lead to a relatively lower risk for incident stroke.

## STUDY STRENGTHS

To our knowledge, this study is the largest cohort with two sequential standardized ETT-CRF assessments at least one year apart. Although there are no widely accepted standards for the classification of CRF based on ETT performance, we followed the age-and-sex-specific methods for CRF standardization proposed in our previous work.^32^ The Veterans Health Administration system provides equal access to health care independent of a patient’s financial status, which minimized the effect of disparities in medical care.^26–28^ This, along with the existence of electronic health records within the Veterans Health Administration system, enables the assessment of history and alterations in health status to be analyzed in detail. ^25, 28^ These factors, coupled with the similar trends in the CRF-stroke association observed when we excluded individuals who developed stroke within three years post hypertension, as well as those who died within the initial 3 years of follow-up, reduce the effects of pre-existing disease on our findings. Thus, the probability of reverse causality was less likely, strengthening the validity of the association between change in CRF and stroke risk.

We recognize that changes in CRF are in part age-related.^33^ Hence, to account for the impact of aging on CRF, we adjusted the model for the time (years) between the two CRF assessments. Thus, our finding of changes in CRF reflecting inverse and proportional changes in the risk of stroke strengthens the concept that CRF-related reduction in the risk of stroke is independent of genetic factors.

## STUDY LIMITATIONS

We acknowledge the epidemiologic nature of the study precludes prove of causation, regardless of the strong associations between CRF or changes in CRF and risk of stroke. We cannot discern whether the lower risk of stroke is exclusively the result of increases in aerobic capacity or physical activities (aerobic and anaerobic). In addition, the results may have been influenced by factors that were beyond the capacity of this study to account; medical treatment or other interventions may have changed during the follow-up, all of which may have affected the findings. We also cannot discern whether relatively low CRF status and reductions in CRF were the outcome of intentional abstinence from physical activity, other lifestyle factors, or subclinical disease which underlies low CRF (reverse causality), despite steps taken to minimize the probability of reverse causality.

Moreover, we could not account for factors that lead to another ETT following at least one year from the initial ETT (mean time between ETTs 6.0±3.9 years) in approximately 23% of the cohort (n=110,576). It is reasonable to assume that the additional ETT was initiated for clinical reasons. A substantially higher rate of subsequent ETTs for individuals in the Least-fit category (referent) would suggest a higher likelihood of cardiac disease in those individuals. If so, the CRF-stroke incidence across the CRF categories that is based on the final ETT may overestimate the favorable impact of CRF on stroke incidence. We found no evidence to support this assumption. To the contrary, in our cohort approximately 23% of patients had a subsequent ETT and the rate of a subsequent ETT across CRF categories was progressively higher with increased CRF (22% for the Least-fit to 24% for the High-fit).

We also acknowledge that our cohort was drawn from ETTs performed across the VA health care system, and while the same protocol was used, some unknown differences in ETT conduct may exist. Lastly, our population (US Veterans referred for ETT) may not reflect the general population.

### Conclusions

The salient message from the present study is that relatively poor CRF was a stronger predictor of stroke than any of the traditional risk factors examined regardless of age, sex, or race. Furthermore, changes in CRF reflected inverse changes in stroke risk, suggesting that stroke risk can be modulated by CRF.

## Data Availability

The dataset of this study was derived from the ETHOS dataset. Access to this dataset is granted to scientist affiliated with the Veterans Affairs Health Care System.

## CLINICAL PERSPECTIVE

Our findings strongly support that poor cardiorespiratory fitness (CRF) is the strongest predictor of risk for all types of stroke in hypertensive patients regardless of race and age. The CRF-stroke risk association is inverse and graded. Additionally, change in CRF is inversely related to changes in stroke risk, suggesting that stroke risk can be modulated by improved CRF. Significantly lower risk rates were observed with peak CRF of approximately 5.0, 6.0 and 7.0 METs for ages ≥70, 60-69 and <60 years, respectively. These MET levels are achievable by most middle-aged and older individuals by meeting current guidelines that advocate ≥150 minutes per week of moderate or 75 minutes of higher intensity physical activity. Accordingly, individuals should be encouraged to initiate and maintain a physically active lifestyle consisting of moderate-intensity activities (brisk walking or similar activities) at any age. Such programs are likely to improve exercise capacity and lower the risk of stroke.

